# Abnormal synergies and associated reactions post-hemiparetic stroke reflect the neuroanatomy of brainstem motor pathways

**DOI:** 10.1101/2022.04.25.22273876

**Authors:** Laura M. McPherson, Julius P. A. Dewald

## Abstract

Individuals with moderate-to-severe post-stroke hemiparesis have difficulty controlling proximal and distal joints of the arm independently because they are constrained to stereotypical movement patterns called the flexion and extension synergies. Over the last three decades, we and others have quantitatively characterized these patterns and have provided evidence that they emerge because of an increased influence of diffusely-projecting brainstem motor pathways following stroke-induced damage to corticospinal and corticobulbar pathways. In our recent work that has focused on how they influence post-stroke hand function, we observed three notable aspects of synergy expression that we have never studied systematically: (1) paretic wrist and finger muscles were often activated maximally while individuals contracted muscles at a different joint, not during a maximal voluntary contraction of the wrist and finger muscles themselves; (2) there were differences in the magnitude of synergy expression when elicited via contraction of proximal muscles vs. distal muscles; (3) there was consistent movement resembling flexion or extension synergy patterns in the paretic limb during maximal efforts with the non-paretic limb (a phenomenon described clinically as an associated reaction), and the strength of these movement seemed to differ based on which muscles in the non-paretic limb were being activated. In the current study, we investigated the above behaviors systematically during maximal isometric contractions of shoulder, elbow and wrist/finger muscles, specifically focusing on differences between proximal vs. distal joints and flexor vs. extensor muscles. Our overall hypothesis is that the muscle-dependent nature of the behaviors we have observed is consistent with how the muscles are impacted by corticofugal damage and the upregulation of brainstem motor pathways that results. That is, we expected that our findings would reflect the fact that the greatest proportion of descending neural control comes from the corticospinal tract for distal muscles and from brainstem motor pathways for proximal muscles. We further expected that findings would reflect the fact that the reticulospinal tract, thought to underlie the flexion synergy, has bilateral effects in the upper limbs and favors facilitation of flexor muscles on the ipsilateral side. Supporting this hypothesis, we found that for some participants, joint torque and muscle activation generated during maximal voluntary contractions were lower than during maximal synergy-induced contractions. This was more prevalent and more severe in magnitude at the wrist and fingers than at the shoulder and elbow. We also found that synergy-driven contractions were strongest when elicited via proximal joints and weakest when elicited via distal joints. Finally, we found that associated reactions in the paretic wrist/finger flexors were stronger than those of other paretic muscles and were the only ones whose response depended on whether the contralateral contraction was at a proximal or distal joint. Our results provide indirect evidence for how an increased reliance on brainstem motor pathways post-stroke contributes to abnormal motor behaviors and demonstrate the need to examine whole-limb behavior when studying or seeking to rehabilitate the paretic upper limb.

## 1 Introduction

Stereotypical movement patterns that emerge in the upper limb of individuals with moderate-to-severe post-stroke hemiplegia present a substantial barrier to completing functional tasks because they interfere with the ability to control proximal and distal joints independently. These obligatory movement patterns are described clinically as the flexion synergy (shoulder abduction coupled with elbow, wrist, and finger flexion) and the extension synergy (shoulder adduction coupled with elbow extension, wrist flexion or extension, and finger flexion) (Brunnstrom 1970; Dewald and Beer 2001; Lan et al. 2017; McPherson and Dewald 2019; Miller and Dewald 2012). They emerge as a result of an increased influence of diffusely-projecting brainstem motor pathways following stroke-induced damage to the corticospinal pathway (Karbasforoushan et al. 2019; McPherson et al. 2018a, 2018b; Owen et al. 2017).

Over the last decade, we have extensively characterized the flexion and extension synergies at the shoulder, elbow, wrist, and fingers (Lan et al. 2017; McPherson and Dewald 2019; Miller and Dewald 2012), extending the laboratory’s previous work that focused on the proximal manifestation of the synergies at the shoulder and elbow joints (Dewald et al. 1995, 2001; Dewald and Beer 2001; Ellis et al. 2007, 2012; Sukal et al. 2007). Over the course of these experiments, we have observed three notable aspects of synergy expression that we have never studied systematically. First, we noticed that paretic wrist and finger muscles were often activated maximally while individuals contracted muscles at a different joint, not during a maximal voluntary contraction of the wrist and finger muscles themselves, as is typical. Second, we noticed differences in the magnitude of synergy expression when it was elicited via contraction of proximal muscles vs. distal muscles. Third, we noticed consistent movement resembling flexion or extension synergy patterns in the paretic limb during maximal efforts with the non-paretic limb (a phenomenon described clinically as an associated reaction). The strength of these associated reactions seemed to differ based on which muscles in the non-paretic limb were being activated. Therefore, the goal of this study was to quantify and characterize the above behaviors systematically, specifically focusing on differences between proximal vs. distal joints and flexor vs. extensor muscles.

Our overall hypothesis is that the muscle-dependent nature of the behaviors we have observed is consistent with the ways in which the muscles are impacted by the increased utilization and upregulation of brainstem motor pathways that occurs following stroke-induced corticospinal and corticobulbar tract damage. All upper limb muscles are controlled by both the precise, sophisticated lateral corticospinal system and the diffusely-projecting, comparatively-more-crude brainstem motor system (e.g., reticulospinal, vestibulospinal pathways). However, these two motor systems have different contributions to neural control of proximal vs. distal muscles. Brainstem motor pathways have the strongest projections to proximal muscles (Davidson & Buford, 2006; Lawrence & Kuypers, 1968a, 1968b), which is in line with their role in postural stability. Conversely, the corticospinal pathway has the strongest and most frequent projections to distal muscles (McKiernan et al., 1998), which is in line with their role as the predominate muscles for fine motor control. In addition, the reticulospinal pathway, which is the brainstem pathway thought to underlie the flexion synergy, has bilateral effects in the upper limbs and favors facilitation of flexor muscles on the ipsilateral side (Davidson et al. 2007; Davidson and Buford 2004, 2006; Herbert et al. 2010; Schepens and Drew 2006). Following corticospinal and corticobulbar damage, activity of brainstem pathways may be inadequately balanced due to the reduced activity of the corticospinal tract and/or a loss of oligosynaptic inhibitory cortico-reticular connections (Fisher et al. 2021). As a result, the way that muscles are activated may reflect characteristics of brainstem pathways.

Based on this overall hypothesis, our study has the following aims and specific predictions. The first aim of the study was to determine which paretic upper limb muscles are activated maximally during contractions of muscles at other joints (i.e., during elicitation of the flexion and extension synergies) rather than during voluntary contractions of the muscles themselves. We predicted that maximal activation of proximal muscles (i.e., those of the shoulder and elbow) would be achieved through voluntary contractions but that maximal activation for the most distal muscles (i.e., those of the wrist and fingers) would occur during synergy-driven contractions.

The second aim of the study was to determine whether the magnitude of flexion and extension synergy expression differs when elicited via maximal contractions of proximal vs. distal muscles in the paretic arm. Similarly, the third aim of the study was to determine whether the magnitude of associated reactions differs when elicited via maximal contractions of proximal vs. distal muscles of the *contralateral* (non-paretic) arm. Because proximal muscles are more heavily innervated by brainstem pathways than are distal muscles, we predicted that activation of proximal muscles would result in stronger synergy expression and associated reactions compared with activation of distal muscles.

Our findings are consistent with our predictions. For some participants, joint torque and muscle activation generated during maximal voluntary contractions were lower than during maximal synergy-induced contractions. This was more prevalent and more severe in magnitude at the wrist and fingers than at the shoulder and elbow. Synergy-driven contractions were strongest when elicited via proximal joints and weakest when elicited via distal joints. Associated reactions in the paretic wrist/finger flexors were stronger than those of other paretic muscles and were the only ones whose response depended on whether the contralateral contraction was at a proximal or distal joint.

Portions of the data have been reported in abstract (Miller and Dewald 2014) and dissertation (Miller 2014) form.

## 2 Methods

### 2.1 Participants

Individuals with chronic hemiparetic stroke (>1 year prior) were recruited through a departmental research database. Participation required enough passive range of motion at the shoulder, elbow, wrist, and fingers to be placed comfortably in the isometric testing setup (described in the following subsection). A physical therapist performed a clinical exam on potential participants. Clinical motor deficits that were acceptable for inclusion in the study were those consistent with cortical or sub-cortical lesions (e.g., unilateral hemiparesis with non-cerebellar, non-brainstem clinical signs). We could obtain specific lesion locations for 9 of the 12 participants from either their medical records, or when available, a computed tomography scan and/or a T1-weighted magnetic resonance imaging scan.

For scans that had not been interpreted by a radiologist, research personnel with training in neuroanatomy identified lesion locations. Exclusion could result from of any one of the following four conditions: first, an upper extremity Fugl-Meyer Motor Assessment (FMA) (Fugl-Meyer et al. 1975) score outside of the 10–44 range (0–9 indicating near paralysis and 45–66 indicating mild impairment); second, a Chedoke-McMaster Stroke Assessment hand portion (CMSAh) (Gowland et al. 1995) score greater than 5 (indicating mild impairment); third, significant impairment of vision or upper extremity tactile somatosensation; or fourth, the recent use of Botox^®^ or Dysport^®^ in the paretic upper limb.

Twelve individuals with chronic post-stroke hemiparesis met all inclusion criteria and completed the study (three females, nine males; mean age ± SD: 59.0 ± 6.2 years, range 47-70; mean ± time post-stroke 10.3 ± 5.9 years; range: 3.5– 26.6; Table 1). Participants exhibited severe-to-moderate upper limb motor impairment according to the FMA with scores ranging from 13 to 30 of 66 possible (mean: 22.7). They also exhibited severe-to-moderate hand motor impairment according to the CMSAh with score ranging from 2 to 4 of a possible 7 (mean: 3.0). Seven participants had right-sided hemiparesis and five had left-sided hemiparesis.

**Table 1.** Demographics of the participants with chronic hemiparetic stroke

| Participant | Sex | Affected/<br>Dominant | Lesion Location | Yrs post-stroke | FMA | CMSAh |
| --- | --- | --- | --- | --- | --- | --- |
| 1 | F | R/R | BG, TH, IC | 26.6 | 20 | 3 |
| 2 | M | L/L | N/A | 11.4 | 24 | 2 |
| 3 | M | R/R | BG, IC, CFL, SFL, IN | 4.5 | 13 | 3 |
| 4 | F | R/R | BG, IC, TH, HC | 5.3 | 30 | 3 |
| 5 | M | L/L | IC | 3.8 | 25 | 4 |
| 6 | M | L/R | BG, IC | 6.5 | 24 | 3 |
| 7 | F | L/R | N/A | 9.0 | 15 | 3 |
| 8 | M | R/L | BG, TH, IC, CTL | 17.0 | 19 | 2 |
| 9 | M | R/L | N/A | 25.5 | 26 | 3 |
| 10 | M | L/L | CPL, CFL, CTL | 5.6 | 31 | 3 |
| 11 | M | R/L | IC | 5.1 | 20 | 4 |
| 12 | M | R/R | IC | 3.5 | 29 | 3 |
| Mean (SD) |  |  |  | 10.3 (8.3) | 22.7 (5.9) | 3.0 (0.6) |

FMA = Upper extremity score of the Fugl-Meyer Motor Assessment (max = 66); CMSAh = Hand portion of the Chedoke-McMaster Stroke Assessment (max = 7); BG = Basal Ganglia; TH = Thalamus; IC = Internal Capsule; CFL = Cortical Frontal Lobe; SFL = Subcortical Frontal Lobe; CPL = Cortical Parietal Lobe; HC = Hippocampus; IN = Insula; N/A = Not Available

Six control participants without known neurological injury (four males, two females; mean age 60.6 years) were included for comparison with the non-paretic limb of participants with stroke. All participants gave informed consent for participation in the study, which was approved by the Institutional Review Board of Northwestern University.

### 2.2 Experimental setup and data collection

The experimental protocol was conducted in a testing device capable of measuring isometric shoulder, elbow, wrist, and finger (metacarpophalangeal) joint torques simultaneously (McPherson and Dewald 2019; Miller et al. 2014). Participants were seated in an experimental chair (Biodex, Inc.) with shoulder/waist restraints to prevent shoulder girdle and trunk motion. The tested forearm was placed in a fiberglass cast to interface the arm rigidly with a six degree-of-freedom load cell (JR3, model 45E15A) through a Delrin ring. The wrist and fingers were placed in a custom Wrist and Finger Torque Sensor (WFTS) (Stienen et al. 2011). The arm was positioned in 75° shoulder abduction, 40° horizontal adduction, 90° elbow flexion, 15° pronation, and 0° wrist flexion/extension. Paretic fingers were positioned at 15° finger flexion to accommodate range of motion restrictions. The non-paretic and control fingers had to be positioned at 0° finger flexion/extension because the increased strength of these groups slightly deformed the WFTS attachment bracket such that it would interfere with the isometric device if positioned at 15° of finger flexion. The contralateral (non-tested) arm rested comfortably at each participant’s side. A computer monitor displayed real-time visual feedback of joint torque data.

Electromyographic (EMG) activity was recorded using active differential surface electrodes with a 1-cm interelectrode distance (16-channel Bagnoli EMG System; Delsys, Inc.; 1000x gain, 20–450 Hz bandpass). On the tested arm, electrodes were placed over the following muscles according to the landmarks described by Perotto and Delagi (1994): anterior deltoid (DELT), sternocostal head of the pectoralis major (PEC), biceps brachii (BIC), lateral head of the triceps brachii (TRI), extrinsic wrist and finger flexors (flexor carpi radialis (FCR), flexor digitorum profundus (FDP)), an intrinsic finger flexor (first dorsal interosseous (FDI)), extrinsic wrist and finger extensors (extensor carpi radialis (ECR), extensor digitorum communis (EDC)), and two thumb muscles (flexor pollicis brevis (FPB) and extensor pollicis longus (EPL)). In addition, electrodes were placed on the DELT, BIC, TRI, FCR, and ECR muscles of the contralateral arm. A signal conditioner (Frequency Devices, Model 9064) filtered (8th order Butterworth low-pass filter, 500 Hz) and amplified EMG and wrist and finger torque data before digitization at a sampling frequency of 1 kHz. A handgrip dynamometer collected maximum grip forces used for descriptive purposes.

### 2.3 Experimental protocol

#### Ipsilateral contractions

Each participant’s maximum voluntary torques (MVTs) and corresponding maximum voluntary muscle contractions (MVC) were measured in the tested arm during isometric torque generation in the following directions: shoulder abduction (SABD), shoulder adduction (SADD), elbow flexion (EF), elbow extension (EE), wrist flexion (WF), wrist extension (WE), finger flexion (FF), finger extension (FE), thumb flexion (for FPB MVC only) and thumb extension (for EPL MVC only). MVT directions were randomized, and trials within a direction were repeated until three trials with peak torque within 85% of the maximum torque value were obtained. If the last trial produced the largest peak torque, additional trials were collected. Participants were given vigorous verbal encouragement throughout MVT trials.

Visual feedback of torque in the target direction was shown except for during the WE and FE tasks while testing the paretic limb. Because most participants with stroke had little-to-no voluntary WE or FE on the paretic side, efforts to produce these movements often resulted in flexion (a phenomenon described previously (Kamper et al. 2003; McPherson and Dewald 2019; Miller and Dewald 2012)). Therefore, no visual feedback was given for these directions to ensure participants’ maximal effort.

#### Contralateral contractions

Maximum voluntary efforts in SABD, EF, EE, WF and WE directions were performed for the arm contralateral to the tested arm. A physical therapist manually resisted the contralateral arm during these efforts, and the response of the tested arm was measured in the isometric testing device.

### 2.4 Data analysis

All data analysis was performed using custom MATLAB software. A Jacobian-based algorithm was used to convert forces and moments collected from the six degree-of-freedom load cell attached at the forearm into shoulder and elbow joint torques. Torque and full-wave rectified EMG data were smoothed using an acausal one-sided moving average filter of window length 250 ms, baseline corrected so that any muscle tone at rest would not factor into subsequent analyses and normalized to the largest value obtained over the experiment, but only during voluntary activation of the tested arm (not during contralateral torque generation). This normalization value was chosen instead of the maximum value during a voluntary contraction, because paretic limb voluntary wrist and finger torque and EMG values were often very small in comparison to values generated during other MVT directions, resulting in inflated values when normalized.

For each MVT trial in the tested arm, maximal torque in the primary (intended) direction was determined. Secondary torques (i.e., those in degrees-of-freedom other than the primary direction) at the time of maximal primary torque were collected, as were EMG values at 50 ms preceding the maximal value, to account for an estimate of the electromechanical delay inherent to skeletal muscle (Cavanagh and Komi 1979).

To compare muscle activation during voluntary vs. synergy-driven contractions, we calculated a “voluntary activation deficit” as follows. We divided the maximal EMG value obtained during maximal voluntary contractions (i.e., when the muscle performed as an agonist, e.g, during finger flexion for FCR and FDP) by the maximal EMG value obtained during maximal contractions in all torque directions. We then subtracted this ratio from a value of 1 so that low values would indicate a small deficit in activation during voluntary contractions and high values would indicate a large deficit in activation during voluntary contractions, and it was multiplied by 100 to achieve a percentage. A value of 0% indicates a muscle’s largest EMG value occurred during a voluntary contraction, and a value of 90% indicates a muscle’s EMG value during voluntary contraction is 10% of the EMG value obtained during the largest synergy-driven contraction. We made the same calculation with torque data to compute the “voluntary strength deficit.”

To calculate the magnitude of synergy expression that resulted from each primary torque direction, mean synergy-driven torque was calculated by averaging the magnitude of all secondary torques.

For each MVT trial performed in the contralateral arm, maximal EMG of the agonist muscle was determined and EMG from the tested arm at that time point was collected.

### 2.5 Statistical analysis

For statistical analyses, *p* ≤ 0.05 was used to determine significance. Cases where *p*-values were greater than 0.05 but less than 0.10 are presented. Values in the text are presented as mean ± SEM unless otherwise specified; values in Table 2 are presented as mean ± SD.

**Table 2.** Maximum voluntary strength measurements

|  | *Maximal voluntary strength in primary direction* |  |  | *Participants in paretic group with synergy-driven strength greater than voluntary strength* |  |  |
| --- | --- | --- | --- | --- | --- | --- |
|  | *Control* | *Non-Paretic* | *Paretic* | *Number* | *Mean(range) percent increase from voluntary to synergy-driven* | *Direction eliciting maximum torque* |
| SABD | 50.1 ± 21.1 | 37.3 ± 17.5 | * 24.1 ± 13.2 | 0 | — | — |
| SADD | 67.8 ± 21.8 | 49.0 ± 23.2 | * 24.9 ± 11.6 | 2 | 18.6 (4.0 – 33.3) | EE x 2 |
| EF | 58.8 ± 18.6 | 51.3 ± 21.4 | * 24.0 ± 11.8 | 3 | 7.1 (1.0 – 16.5) | SABD x 3 |
| EE | 47.5 ± 16.0 | 38.1 ± 15.3 | * 18.5 ± 8.4 | 1 | 11.7 | SADD |
| WF | 18.5 ± 4.5 | 15.1 ± 5.6 | * 3.9 ± 1.8 | 5 | 48.5 (9.2 – 102.1) | SABD x 3, EF x 2 |
| WE | 8.6 ± 2.1 | 6.8 ± 2.2 | — | — | — | — |
| FF | 11.3 ± 2.7 | 7.7 ± 3.3 | * 3.0 ± 1.5 | 5 | 37.6 (9.0 – 103) | SABD x 1, SADD x 1, EF x 3 |
| FE | 2.1 ± 0.5 | 2.1 ± 0.7 | — | — | — | — |
| Grip | 417.3 ± 108.9 | 376.8 ± 115.7 | * 62.0 ± 28.0 | — | — | — |

Values are group mean ± standard deviation. Strength data are displayed in Newton-meters, except for grip strength which is displayed in Newtons. An asterisk indicates a significant difference between paretic and non-paretic groups, and $p$ -values are reported in the text.

To compare maximal voluntary strength between paretic and non-paretic limbs, we used a 2 × 7 linear mixed effects model to test effects of the fixed, repeated factors of limb (paretic, non-paretic) and primary torque direction (SABD, SADD, EF, EE, WF, FF, Grip) as well as the limb-by-primary torque direction interaction on maximum voluntary torque (or force, in the case of Grip strength) (GraphPad Prism, v8). Participant was included as a random factor, and the Greenhouse-Geiser correction was applied. The WE and FE directions were not included in this model because none of the paretic limbs could generate voluntary torque in these directions. To compare maximal voluntary strength between non-paretic and control limbs, we used a 2 × 9 linear mixed effects model to test effects of the fixed factors of limb (non-paretic, control) and primary torque direction (repeated; SABD, SADD, EF, EE, WF, FF, WE, FE, Grip) as well as the limb x primary torque direction interaction on maximum voluntary torque/force (GraphPad Prism, v8). Participant was included as a random factor, and the Greenhouse-Geiser correction was applied. The effects of interest for both of the above models were the main effect of limb and the limb-by-direction interaction. Planned comparisons on the interactions using Fisher’s least square difference tests determined differences between limbs for each primary torque direction.

To evaluate whether the voluntary strength deficit in torque differed among proximal and distal contraction directions, we used a 1-way repeated measures Friedman test (due to non-normally distributed data per the Shapiro-Wilk test).

To evaluate whether the voluntary activation deficit in EMG differed among proximal and distal contraction directions, we used a 1-way repeated measures linear mixed effects model with random factor of participant (GraphPad Prism, v8). Then, we conducted planned comparisons using an uncorrected Dunn’s test to determine whether voluntary activation deficit values for each muscle differed from those for the deltoid. Out of 156 data points (12 participants x 13 muscles) there were 5 instances of missing data due to poor signal quality: one from the ECR, two from the FPB, and 2 from the EPL.

To determine differences in secondary torque generation between paretic and non-paretic limbs for each primary torque direction, we used a 2 × 8 repeated measures ANOVA (GraphPad Prism, v8) to test the main effect of limb and the limb-by primary torque direction interaction on joint torque. We used planned comparisons on the interaction using Fisher’s least square difference tests to determine differences between limbs for each torque direction.

To determine whether there are differences in the strength of synergy expression for proximal-to-distal and distal-to-proximal joint combinations, we used a 1-way ANOVA to test for differences among joint combinations for joints that elicited the flexion synergy. Then, we conducted planned comparisons using Fisher’s least square difference tests among the salient joint combinations (see Results). We repeated these tests with data from joints the elicited the extension synergy. We also used a 1-way ANOVA to test for differences in the strength of overall synergy expression for each primary torque direction, followed by planned comparisons that compared data from each primary torque direction with that of the SABD direction.

To examine the effect of contralateral muscle contractions on the tested arm, we averaged EMG data from the three wrist and finger flexors (FCR, FDP, FDI) to establish an EMG value for the wrist/finger flexor muscle group as a whole for brevity. In the same way, we established EMG values for the wrist/finger extensor muscle group by averaging EMG values from the ECR and EDC. To test differences between paretic and non-paretic limbs for each muscle group (wrist/finger flexors, wrist/finger extensors, biceps, triceps), our model included fixed factors of limb, contraction direction, and the limb-by-contraction direction interaction (R, version 3.6.3 with lme4 package (Bates et al. 2015)). A random intercept was included, clustered by participants. To test differences in flexor and extensor muscle groups within the paretic limb, our model included fixed factors of muscle group, contraction direction, and the muscle group-by-contraction direction interaction. A random intercept and random slopes of muscle group and torque direction were included (clustered by participants). Estimation of fixed and random effects for all models used Restricted Maximum Likelihood Estimation.

## 3 Results

Maximum voluntary strength measurements from the paretic, non-paretic, and control limbs are shown in Table 2. The paretic limb was significantly weaker than the non-paretic limb in all directions (main effect of limb: *F*(1, 11) = 160.3, *p* < 0.0001; limb-by-direction interaction: *F*(1.1, 11.9) = 117.8, *p* < 0.0001; SABD: t(11) = 3.9, *p* = 0.002; SADD: t(11) = 5.6, *p* = 0.0002; EF: t(11) = 6.7, *p* < 0.0001; EE: t(11) = 3.9, *p* = 0.003; WF: t(11) = 7.9, *p* < 0.0001; FF: t(11) = 6.6, *p* < 0.0001; Grip: t(11) = 11.7, *p* < 0.0001), and none of the paretic limbs could generate appreciable wrist or finger extension torque. There were no overall differences between the non-paretic and control limbs (main effect of limb: *F*(1,16) = 1.8, *p* = 0.20; limb-by-direction interaction: *F*(8, 128) = 0.49, *p* = 0.86).

### 3.1 Voluntary vs. synergy-driven maximal torque and muscle activation at proximal vs. distal joints

Eight of the twelve participants generated more paretic torque during a synergy-driven contraction than during a voluntary contraction for at least one joint (Table 2, Figure 1A). SABD was the only direction for which all participants generated maximal torque during the voluntary contraction. For the remaining contraction directions, the numbers of participants with maximal torque during the voluntary contraction was fewer at the wrist and finger joints than shoulder and elbow joints (SADD: 10, EF: 9, EE: 11, WF: 5, FF: 5) (significant effect of contraction direction on voluntary strength deficit (*p* = 0.029). For the participants whose maximum torque was achieved via synergy-driven rather than voluntary efforts, the discrepancy between the voluntary and synergy-driven activation was substantially greater for the wrist and finger joints, with the voluntary strength deficit values averaging 14.4%, 6.2%, 10.5%, 29.5%, 23.3% for the SADD, EF, EE, WF, and FF directions, respectively. The combinations of joints were within the flexion and extension synergy patterns (e.g., maximal EF torque produced during SABD and maximal EE torque produced during SADD; Table 2).

**Figure 1.**
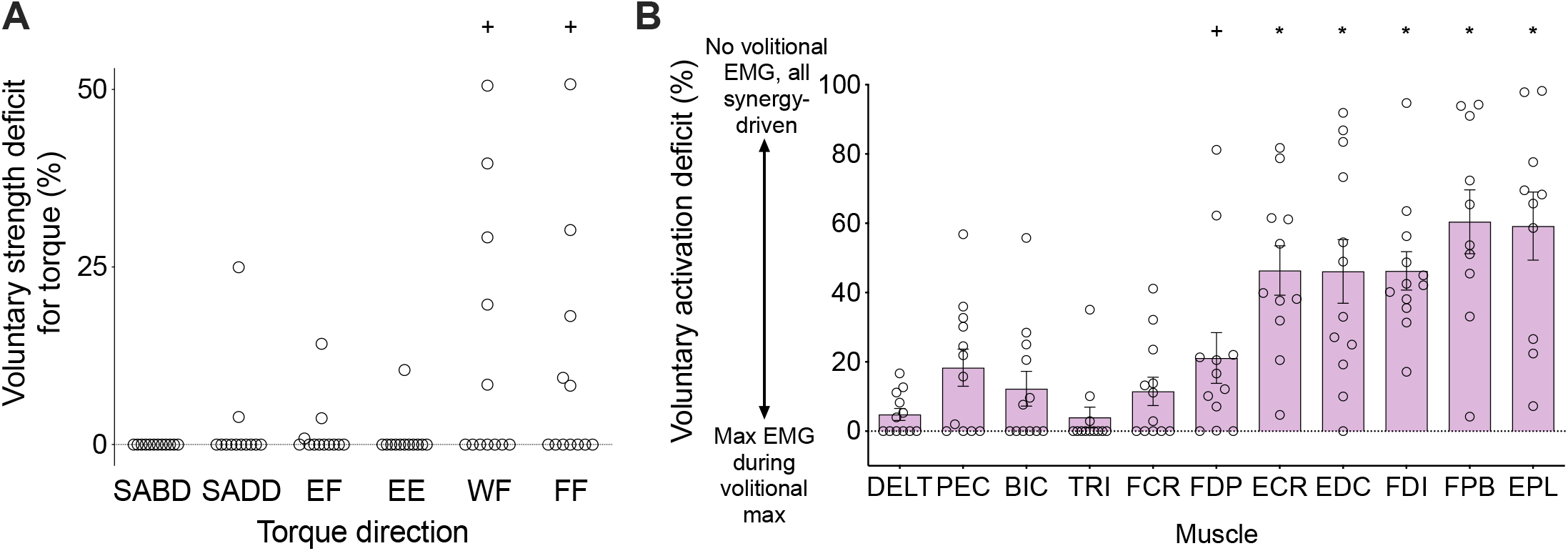
Voluntary strength deficit in torque (A) and voluntary activation deficit in EMG (B) for the paretic limbs. A value of 0 indicates that maximal torque/EMG was obtained during a contraction in the intended/agonist direction, and non-zero values indicate that maximal torque/EMG was obtained during a contraction in a different direction (i.e., synergy-driven). The higher the value, the larger the discrepancy between the voluntary and synergy-driven values. Asterisks indicate significant differences between values for a particular torque direction/muscle and SABD/deltoid at *p* ≤ 0.05 (*), *p* ≤ 0.10 (**+**). Exact *p*-values are presented in the text.

Similarly, the voluntary activation deficit differed among the 13 muscles (Figure 1B) (significant effect of muscle (*F*(10,105) = 11.6, *p* < 0.0001). It was higher for wrist/finger extensors (*ECR*: 46.1%, *t*(105) = 4.6, *p* < 0.0001; *EDC*: 46.1%, *t*(105) = 4.7, *p* < 0.0001) and intrinsic hand muscles (*FDI*: 46.3%, *t*(105) = 4.8, *p* < 0.0001; *FPB*: 60.4%, *t*(105) = 6.1, *p* < 0.0001; *EPL*: 59.2%, *t*(105) = 6.0, *p* < 0.0001) compared with DELT (4.8%). Values for the remaining muscles, including the wrist/finger flexors, were not significantly different from those of the deltoid (although the FDP was significantly higher at the *p* < 0.075 level) (*PEC*: 18.3%, *t*(105) = 1.6, *p* = 0.12; *BIC*: 12.3%, *t*(105) = 0.9, *p* = 0.40; *TRI*: 4.0%, *t*(105) = 0.1, *p* = 0.92; *FCR*: 11.5%, *t*(105) = 0.8, *p* = 0.44; *FDP*: 21.4%, *t*(105) = 1.9, *p* = 0.06).

### 3.2 Proximal vs. distal elicitation of the flexion and extension synergies

Figure 2A shows group mean SABD/SADD, EF/EE, WF/WE, and FF/FE secondary torques produced during the generation of each primary torque direction. Secondary torques of the paretic limb differed from those of the non-paretic limb for all eight primary torque directions (significant limb-by-secondary torque direction interactions: SABD: *F*(3, 33) = 15.4, *p* < 0.0001; SADD: *F*(3, 33) = 17.5, *p* < 0.0001; EF: *F*(3, 33) = 17.7, *p* < 0.0001; EE: *F*(3, 33) = 31.0, *p* < 0.0001; WF: *F*(3, 33) = 5.3, *p* = 0.004; WE: *F*(3, 33) = 47.1, *p* < 0.0001; FF: *F*(3, 33) = 5.3, *p* = 0.004; FE: *F*(3, 33) = 94.8, *p* < 0.0001). Statistically significant planned comparisons on the interaction testing differences between limbs for each secondary torque direction are shown with asterisks in Figure 2A, and *p-*values ranged from < 0.0001 to 0.01.

**Figure 2.**
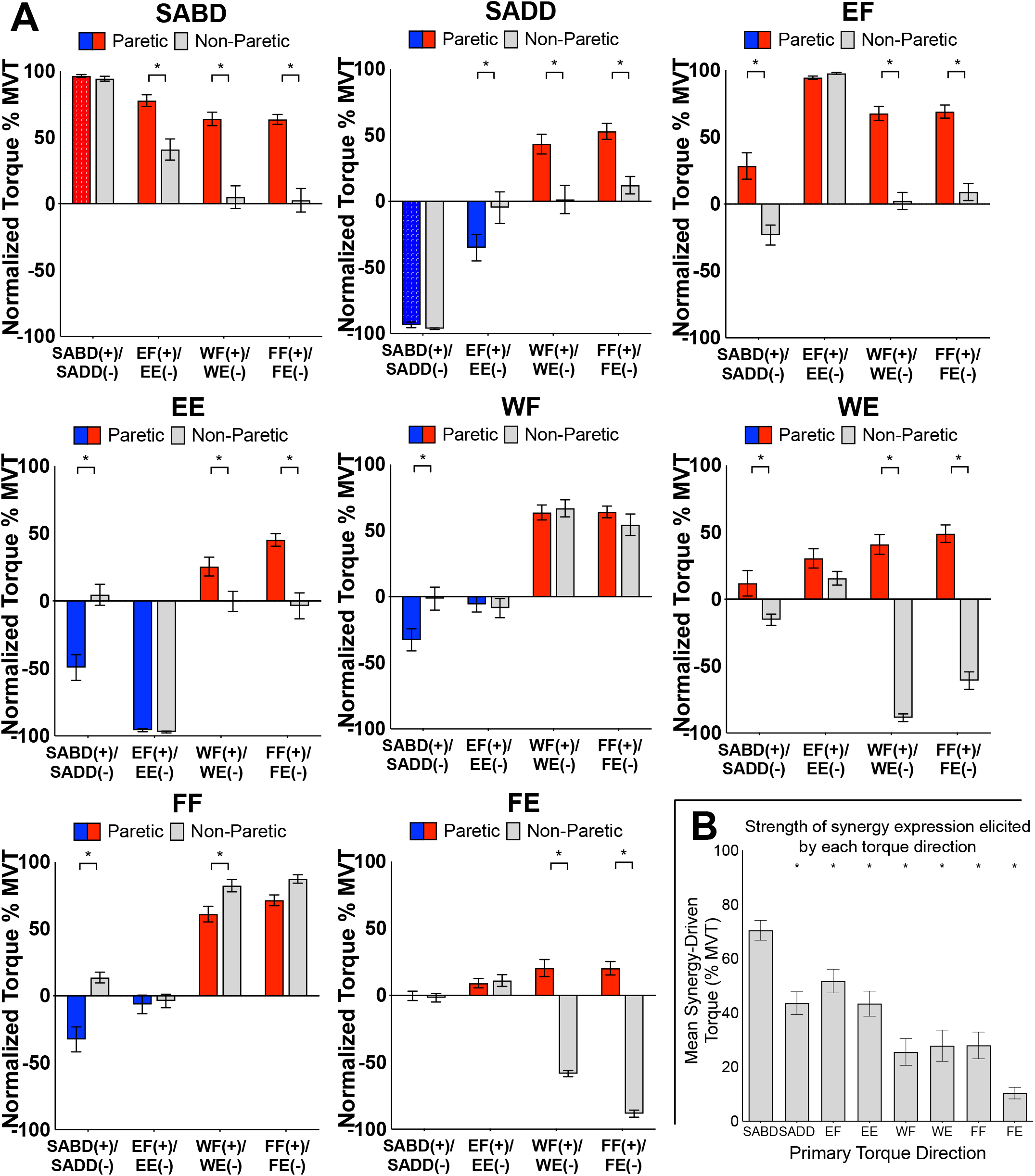
**A**. Group mean ± SEM shoulder, elbow, wrist and finger joint torques produced by the paretic (red/blue) and non-paretic (grey) limbs during generation of SABD, SADD, EF, EE, WF, WE, FF, and FE MVT. Red bars highlight SABD and flexion directions, and blue bars highlight SADD and extension directions for the paretic limb. Asterisks indicate significant differences between paretic and non-paretic limbs (*p-*values range from < 0.0001 to 0.01). **B**. The strength of synergy-driven torque when elicited by each primary torque direction. Asterisks indicate a significantly lower value compared with the synergy-driven torque for SABD (*p-*values range from < 0.0001 to 0.02; exact values are reported in the text).

The overall impact of each primary torque direction on other joints (i.e., the strength of synergy expression elicited by that joint) was different among primary torque directions (Figure 2B, *F*(7, 77) = 24.7, *p* < 0.0001). Planned comparisons revealed the mean synergy-driven torque for SABD (68.1 ± 3.7% MVT) was significantly higher than that of each of the other primary torque directions (*SADD*: 44.9 ± 4.1% MVT, t(77) = 4.3, *p* < 0.0001; *EF*: 57.0 ± 4.0% MVT, t(77) = 2.1, *p* = 0.04; *EE*: 42.7 ± 3.8% MVT, t(77) = 4.7, *p* < 0.0001; *WF*: 25.6 ± 4.9% MVT, t(77) = 7.9, *p* < 0.0001; *WE*: 27.9 ± 5.8% MVT, t(77) = 7.5, *p* < 0.0001; *FF*: 28.0 ± 4.9% MVT, t(77) = 7.5, *p* < 0.0001; *FE*: 10.4 ± 2.1% MVT, t(77) = 10.8, *p* < 0.0001).

Generation of maximal SABD and EF by the paretic limb both resulted in secondary torques that were consistent with expression of the flexion synergy (EF, WF, FF during SABD; SABD, WF, FF during EF), and there was a difference among the joint combinations (significant effect of joint combination, *F*(9, 110) = 22.8; *p* < 0.0001) (Figure 3, left panel). The shoulder-to-elbow effect of the flexion synergy was stronger than the elbow-to-shoulder effect. Generation of maximal SABD induced more secondary EF torque (77.7 ± 4.4% EF MVT) than generation of maximal EF induced secondary SABD torque (28.4 ± 9.9% SABD MVT) (t(110) = 5.7, *p* < 0.0001). Generation of SABD and EF produced similar amounts of secondary WF torque (64.0 ± 5.1% and 67.7 ± 5.3% MVT, respectively, t(110) = 0.43, *p* = 0.67) and secondary FF torque (63.5 ± 3.7% and 69.1 ± 4.9% MVT, respectively, t(110) = 0.64, *p* = 0.52).

**Figure 3.**
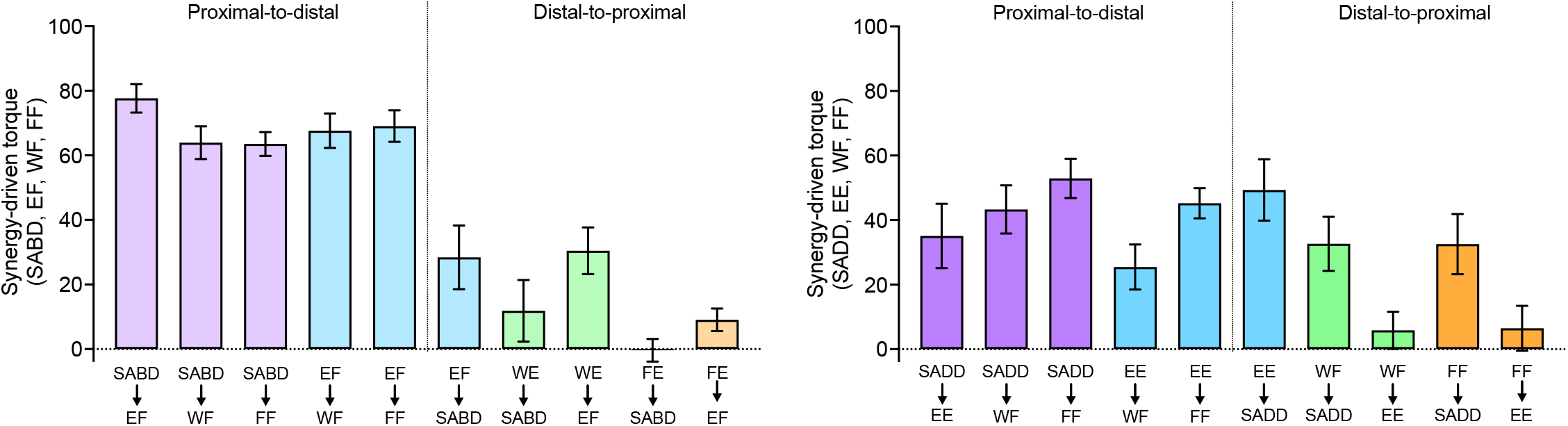
The strength of proximal-to-distal vs. distal-to-proximal elicitation of the flexion and extension synergies. Group mean ± SEM synergy-driven torques (the same as in Fig. 3) are shown for each joint combination and direction, as indicated on the x-axis. The top row of labels is the primary torque direction, and the bottom row of labels is the secondary torque direction. Data from primary torque directions that elicited the flexion synergy are shown in the plot on the left, and data from primary torque directions that elicited the extension synergy are shown on the right. Note that attempts to generate WE and FE (i.e., when they were the primary torque directions) instead resulted in the production of WF and FF, respectively.

During the intended generation of maximal WE and FE torques, the paretic limb group instead produced torque in the WF and FF directions, as we anticipated based on previous literature. Efforts to produce WE torque elicited the flexion synergy in the paretic limb, as evidenced by SABD and EF secondary torques that were small to moderate in magnitude (11.9 ± 9.5% and 30.5% ± 7.2% MVT, respectively). When FE was the primary torque direction, however, virtually no torque as produced at the shoulder for either limb group, and the amount of EF torque was similar between groups. Similar to the shoulder and elbow comparisons, proximal-to-distal elicitation of the flexion synergy led to stronger secondary torques than distal-to-proximal elicitation (SABD led to stronger WF (64.0%) than WE led to SABD (11.9%), (t(110) = 6.0; *p* < 0.0001). The same pattern was seen between the elbow and wrist and elbow and fingers. Maximal EF torque led to greater secondary WF torque (67.7 ± 5.3% MVT) than maximal WE led to secondary EF torque (30.5 ± 7.2% MVT (t(110) = 4.3, *p* < 0.0001), and it led to greater secondary FF torque (69.1 ± 4.9% MVT) than maximal FE led to secondary EF torque (9.1 ± 3.5% MVT, (t(110) = 6.9, *p* < 0.0001)).

Generation of SADD and EE demonstrated secondary torques that were consistent with expression of the extension synergy (EE, WF, FF during SADD; SADD, WF, FF during EE), and there was a significant difference among the joint combinations (significant effect of joint combination, *F*(9, 110) = 4.6, *p* < 0.0001) (Figure 3, right panel). Unlike the flexion synergy, however, the magnitude of extension synergy expression elicited via the shoulder was not stronger than that elicited via the elbow. Secondary EE torque produced during maximal SADD (49.3 ± 9.5% EE MVT) was not different than secondary SADD torque produced during maximal EE (35.1 ± 9.9% SADD MVT) (t(110) = 1.3, *p* = 0.19). Secondary WF torque produced during maximal SADD (42.2 ± 7.1% WF MVT) was not different than that produced during maximal EE (25.5 ± 7.0% WF MVT) (t(110) = 1.6, *p* = 0.10), and secondary FF torque produced during maximal SADD (52.7 ± 6.2% FF MVT) was not different than that produced during maximal EE (46.7 ± 4.4% FF MVT (t(110) = 0.7, *p* = 0.48). During the generation of WF and the generation of FF, the paretic limb produced 32.6 ± 8.4% and 32.6 ± 9.3% MVT of secondary SADD torque, respectively. There was not a difference in extension synergy expression when examining proximal-to-distal vs. distal-to-proximal elicitation between SADD and WF (t(110) = 0.98, *p* = 0.33), and for this comparison between SADD and FF, the difference was significant only at the *p* < 0.075 level (t(110) = 1.87, *p* = 0.06).

There was no appreciable secondary elbow torque produced during WF or FF, although generation of torque in these directions elicited the extension synergy pattern in other degrees of freedom at the shoulder and at the forearm, evidenced by shoulder flexion, shoulder internal rotation and forearm pronation torques that were measured but are not presented in this study. The difference in extension synergy expression when examining proximal-to-distal vs. distal-to-proximal elicitation between EE and WF was significant only at the *p* < 0.075 level (t(110) = 1.81, *p* = 0.07). For this comparison between EE and FF, proximal-to-distal elicitation was greater than distal-to-proximal elicitation (t(110) = 3.57, *p* = 0.0005).

### 3.3 Proximal vs. distal elicitation of contralateral associated reactions

The third aim of the study was to examine the strength of associated reactions in one arm when elicited via the production of maximal joint torque at proximal compared with distal joints of the contralateral arm. We also examined the relative activation of flexor and extensor muscles at the elbow and wrist/fingers. Figure 4A and 4B show paretic group mean EMG (wrist and finger flexors, wrist and finger extensors, BIC, TRI) and torque (wrist and finger flexion/extension, elbow flexion/extension) during maximal voluntary efforts by the non-paretic limb in SABD, EF, EE, WF, and WE directions. Non-paretic data during maximal voluntary efforts by the paretic limb are shown in Figure 4C and 4D.

**Figure 4.**
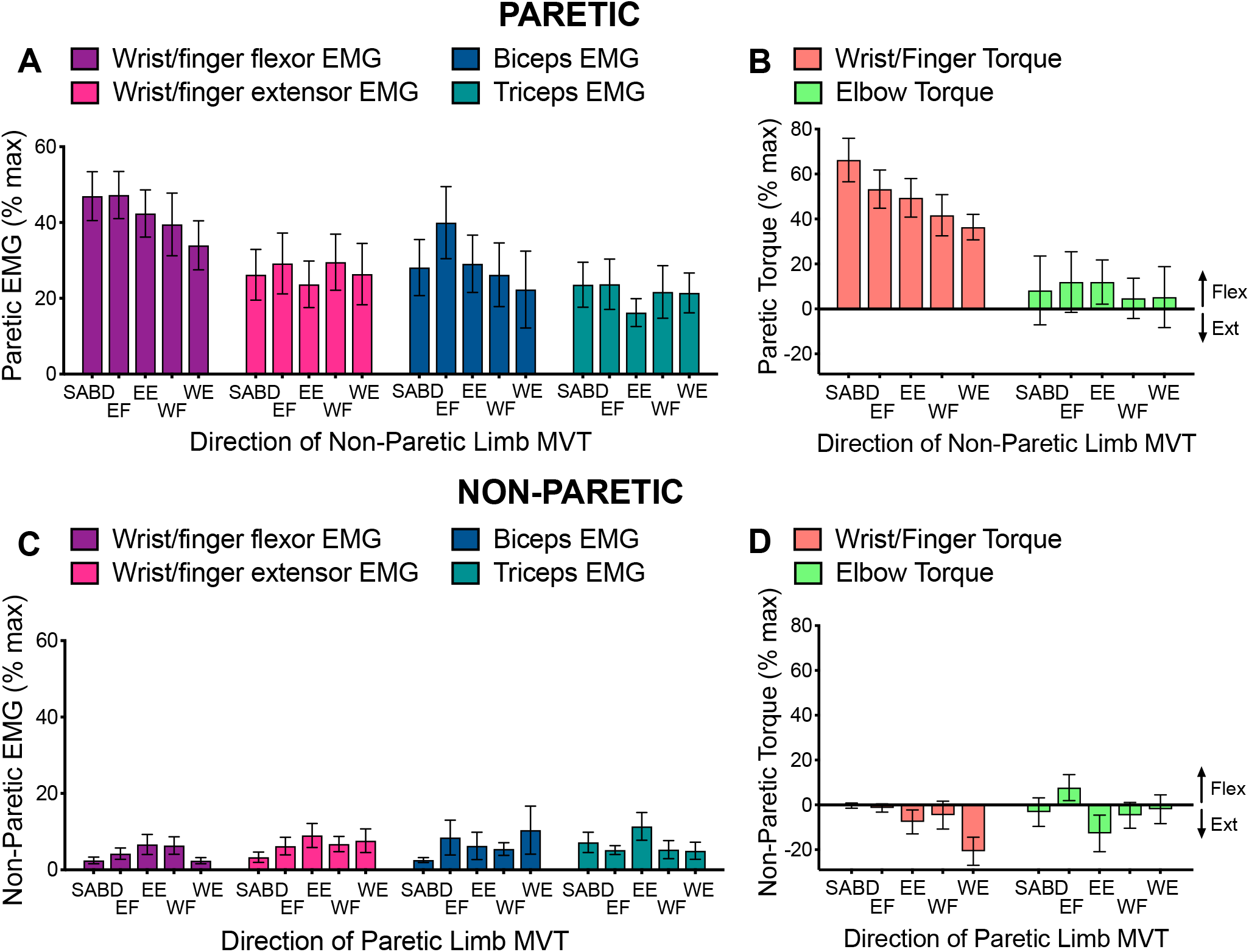
Group mean ± SEM wrist/finger and elbow EMG (**A, C**) and torque (**B, D**) for the paretic (top panel; **A, B**) and non-paretic (bottom panel; **C, D**) limbs during maximal voluntary efforts by the contralateral limb in SABD, EF, EE, WF, and WE directions. Values shown are based on actual data, not the linear mixed effect model estimated values that are presented in the text.

Maximal activation of contralateral muscles produced stronger contractions in the paretic limb than in the non-paretic limb, particularly in the wrist/finger flexors (*wrist/finger flexors*: 41.6 ± 3.1% MVC vs. 3.8 ± 3.1% MVC; significant main effect of limb: *F*(1, 85.0) = 177.0, *p* < 0.0001; *wrist/finger extensors*: 26.3 ± 3.1%, MVC vs. 6.6 ± 3.1% MVC; significant main effect of limb: *F*(1, 86.8) = 38.3, *p* < 0.0001; *BIC*: 29.1 ± 4.0% MVC vs. 5.2 ± 4.0% MVC; significant main effect of limb: *F*(1, 85.9) = 39.5, *p* < 0.0001; *TRI*: 20.8 ± 2.5 vs. 6.4 ± 2.5% MVC; significant main effect of limb: *F*(1, 87.0) = 28.4, *p* < 0.0001). For the non-paretic limb, maximal activation of contralateral muscles (by the paretic limb) produced low levels of co-contraction of flexors and extensors at both the wrist/fingers and elbow.

For some individuals, the paretic wrist/finger EMG and torque values during contralateral torque generation were actually higher than the maximal value that could be produced in the paretic limb during ipsilateral torque generation at any of the joints. The contralateral contractions activated wrist/finger flexor muscles more strongly on average than wrist/finger extensor muscles (41.4 ± 5.4% vs. 25.5 ± 6.2% MVC, significant main effect of muscle group, *F*(1, 10.0) = 9.3, *p* = 0.01; note that these means differ very slightly from those presented in the preceding paragraph because they are marginal means estimated from each statistical model, not the means of the underlying data). In addition, contralateral contractions in the various directions activated the muscle groups differently (significant muscle group-by-contraction direction interaction: *F*(4, 48.3) = 2.9, *p* = 0.03). The wrist/finger flexors demonstrated a decreasing pattern of activation when comparing directions from proximal to distal and the wrist/finger extensors demonstrated an overall consistent pattern of activation among contralateral contraction directions.

For the paretic biceps and triceps, the contralateral contractions produced higher levels of EMG compared with the non-paretic limb, as mentioned above. However, unlike the wrist/finger muscles, the paretic elbow flexor and extensor muscles were activated at similar levels to each other (28.7 ± 7.2% vs. 20.4 ± 4.8% MVC for BIC and TRI, respectively; no significant main effect of muscle (*F*(1, 10.2) = 1.75, *p* = 0.22); no significant muscle-by-contraction direction interaction (*F*(4, 59.1) = 1.59, *p* = 0.19)).

Contralateral contractions resulted in a substantial amount of paretic wrist and finger flexion torque that differed among contraction directions in a decreasing manner from proximal to distal (significant effect of contraction direction (*F*(4, 31.1) = 3.4, *p* = 0.02); group mean values ranging from 66.2 ± 9.7% MVT for the shoulder abduction direction to 36.4 ± 5.7% MVT for the wrist extension direction). For the paretic elbow, contralateral contractions resulted largely in flexion torque (i.e., eliciting the flexion synergy) for all but one participant, who produced maximal levels of elbow extension torque as part of the extension synergy. There was no effect of contraction direction (*F*(4, 32.4) = 0.21, *p* = 0.93). For all participants, paretic elbow flexion torque averaged 8.8 ± 9.8% MVT over contraction directions, with no significant effect of contraction direction. When excluding the participant who exhibited the strong extension synergy, paretic elbow flexion torque averaged 16.7 ± 6.6% MVT.

## 4 Discussion

Our primary findings are that (1) wrist and finger muscles are often activated more strongly during maximal synergy-driven contractions than during maximal voluntary contractions, (2) expression of the flexion and extension synergies is strongest when elicited via proximal rather than distal muscle contractions, and (3) associated reactions in the paretic wrist/finger flexors were stronger than those of other paretic muscles and were the only ones whose response had a proximal to distal decreasing pattern. We interpret our findings as being consistent with an increased influence of brainstem motor pathways, based on the similarly between the effects we saw and the neuroanatomy of this system.

### 4.1 Maximal synergy-driven contractions can be higher than maximal voluntary contractions, particularly for extrinsic wrist/finger extensors and intrinsic hand muscles

We predicted that maximal activation of proximal paretic muscles (i.e., those of the shoulder and elbow) would be achieved through voluntary contractions but that maximal activation for the most distal paretic muscles (i.e., those of the wrist and fingers) would occur during synergy-driven contractions. We reasoned that this finding would be consistent with the ways in which the muscles are impacted by stroke-induced corticospinal and corticobulbar tract damage and the increased reliance on brainstem motor pathways that follows. For example, with corticospinal damage, distal paretic muscles lose more of the neural substrate typically used for voluntary activation compared with proximal muscles, but they can still be activated by brainstem pathways via synergy-driven activation.

Using the voluntary activation deficit to quantify how maximal synergy-driven contractions compare to maximal voluntary contractions, we found that extrinsic wrist/finger extensors (ECR, EDC) and intrinsic hand muscles (FDI, FPB, EPL) had the largest and most frequently occurring increase in synergy-driven activation compared with voluntary activation, which is in line with our prediction. Voluntary activation was only 48% of the synergy-driven activation for those muscles on average, with virtually all participants demonstrating a non-zero voluntary activation deficit. In contrast to our prediction, voluntary activation deficit values for the extrinsic wrist/finger flexors were not statistically different than that for the DELT (although the comparison between FDP and DELT had a *p*-value of 0.06). The difference in the findings between the wrist/finger flexors and the wrist/finger extensors likely reflect the fact that in the intact nervous system, brainstem motor pathways facilitate distal flexors to a greater degree than extensors (Davidson and Buford 2004, 2006), and the strength of this facilitation becomes greater following stroke (Zaaimi et al. 2012). Further, while the strength of corticospinal projections is high to all distal muscles, it is stronger to intrinsic hand muscles and distal extensors compared with distal flexors (Cheney et al. 1991; McKiernan et al. 1998). Thus, following corticospinal damage, it appears that shoulder and elbow muscles and wrist/finger flexors can still be activated voluntarily using brainstem pathways as well as remaining corticospinal resources, whereas wrist/finger extensors and intrinsic hand muscles rely primarily on remaining corticospinal resources.

While the persistence of brainstem pathways following stroke afford the shoulder, the elbow, and the wrist/finger flexors the ability to have some remaining voluntary activation, we must point out that the notable consequence of utilizing predominantly brainstem pathways is the loss of independent joint control that occurs when the flexion and extension synergies are expressed (Karbasforoushan et al. 2019; McPherson et al. 2018b, 2018a; Owen et al. 2017).

### 4.2 Flexion and extension synergy expression is strongest when the synergies are elicited via proximal rather than distal muscle contractions

Because of the aforementioned bias of innervation by brainstem pathways toward proximal muscles, we predicted that activation of these muscles would result in stronger synergy expression when compared with activation of distal muscles. Indeed, our findings support this prediction. When comparing synergy elicitation from one joint to another and vice versa, the proximal-to-distal elicitation was larger than the distal-to-proximal elicitation for every comparison (except for SADD and EE, for which the elicitation was not different between the directions).

When brainstem pathways are activated with the intent to drive shoulder muscles, the elbow and hand are activated as well due to the system’s diffuse multi-joint projections. In the intact nervous system this multi-joint activation may be utilized for postural adjustments and/or to provide multi-joint stability, but the corticospinal tract and its cortico-reticular projections can selectively ‘gate’ or inhibit reticulospinal effects at other joints when they are unwanted (Dyson et al. 2014; Schepens and Drew 2006). Following stroke, however, unwanted effects of brainstem pathways at muscles of other joints are not suppressed, and the flexion and extension synergy patterns emerge. Our findings suggest that the strength of brainstem pathway influence to muscles about one joint determines how strongly the synergy is elicited at muscles of other joints.

### 4.3 Associated reactions are strongest when elicited via proximal rather than distal muscle contractions, but only in the wrist and finger flexors

As expected, we observed strong associated reactions in the paretic limb (unintended activation of paretic muscles that occurred during maximal contractions of the non-paretic limb). We predicted that the associated reactions would be stronger with proximal rather than distal contractions. We found this to be the case for the paretic wrist and fingers. Although strong wrist/finger flexion torque was produced for all contraction directions, it was lower when the contralateral joint was more distal, decreasing by nearly 50% when comparing torque resulting from contralateral shoulder abduction to that of contralateral wrist extension. Nonetheless, there was still an appreciable amount generated during the wrist extension contraction direction. The proximal-distal decreasing pattern in flexion torque across contraction directions was driven by selective decreases in wrist/finger flexor EMG rather than overall decreases in EMG for both flexor and extensor groups. Interestingly, however, paretic elbow torque did not depend on non-paretic contraction direction, evidenced by levels of elbow flexion torque that were consistent across contraction directions and were milder in comparison to that of the wrist/fingers (aside from the one individual who generated maximal levels of elbow *extension* torque).

The presence of the substantial bilateral muscle activity during non-paretic contractions is consistent with the bilateral upper limb projections of the cortico-reticulospinal pathway. It is interesting that the wrist/finger muscles (FCR, FDP, FDI) were the ones that demonstrated the most pronounced activation as well as the dependence on whether the contraction direction was proximal or distal. In studies in non-human primates, the bilateral organization of the reticulospinal tract has been shown to activate muscles as far distal as the wrist (Davidson and Buford 2004, 2006; Herbert et al. 2010), but whether this bilateral organization extends to muscles acting on digits of the hand has not yet been investigated (Baker 2011). While it could be argued that increased activity of the bilaterally projecting reticulospinal tract would also cause associated reactions in the non-paretic limb during paretic limb activation, it is likely that the intact crossed corticospinal tract projecting to the non-paretic limb helps suppress such associated reactions

### 4.4 Implications for clinical research

Results of the study underscore the need to acknowledge whole-limb behavior when examining motor control of the post-stroke upper limb. Studies examining a joint in isolation from the rest of the limb should consider whether results will generalize to functional scenarios when proximal or distal muscles are concurrently activated. Although the current study examines paretic limb behavior during maximal rather than functional efforts, the involuntary coupling between joints via the flexion and extension synergy patterns occurs at submaximal efforts (McPherson and Dewald 2019), including those commensurate with lifting the limb against gravity (Miller and Dewald 2012).

Insight derived from previous studies quantifying flexion and extension synergy expression provided the foundation for a novel physical therapy intervention for reaching based on progressive shoulder abduction loading (Ellis et al. 2008, 2009, 2018) and helped to improve the control of assistive technologies (Makowski et al. 2013, 2014) for the post-stroke upper limb. Results of the current study add to this body of empirical evidence. For example, knowledge of how movement of any of the four paretic upper limb joints (shoulder, elbow, wrist, and finger) elicits the multi-joint synergy patterns could inform the control of a technology that assists the limb differently based on the intended task.

Results suggest that physical therapy interventions using bilateral movements (van Delden et al. 2012) or assistive technologies controlled with the non-paretic limb (Knutson et al. 2009, 2012) may elicit associated reactions in the paretic limb when the amount of effort to the non-paretic limb is high. This may be particularly evident with activation of proximal non-paretic muscles. Additionally, while bilateral training may have important benefits including alterations in intra-cortical inhibition (McCombe Waller and Whitall 2008), it may also upregulate ipsilateral cortico-reticulospinal connections. This would further compound the elicitation of associated reactions during non-paretic limb movement and elicitation of the flexion and extension synergies during paretic limb activation, leading to increased difficulty in controlling joints independently during functional tasks.

### 4.5 Limitations

Several limitations to the study should be considered. First, the sample size was small; however, consistent results were seen in the majority of participants. Second, it should be considered that secondary torques produced during the generation of maximal wrist and finger torques in the paretic limb could be compensatory behaviors (e.g., adducting the shoulder because of the difficulty of wrist/finger flexion or abducting the shoulder than due to the difficulty of wrist/finger extension rather than obligatory synergy-driven shoulder activation). However, if this was the case, the same compensatory behaviors might be expected in non-paretic and control limbs, given that all contractions were maximal efforts and the difficulty between groups would be similar.

Additionally, it is possible that effects of contralateral contractions on the tested limb might have been different if the contralateral arm were in a different position. The effects of paretic upper limb position on ipsilateral reflex behavior (Hoffmann et al. 2009), strength generation (Hoffmann et al. 2011), and extension synergy expression (Ellis et al. 2007) have been previously demonstrated. In particular, the shoulder adduction/elbow extension coupling of the extension synergy was shown to switch to shoulder adduction/elbow flexion when the arm was placed closer to the body as in the current study (Ellis et al. 2007). However, the influence of non-paretic upper limb position on paretic upper limb is unknown.

Finally, the neuroscientific implications drawn from the results are speculative given the behavioral nature of our measurements. However, they are consistent with recent work that has been able to probe the involvement of various neural circuits more directly (Karbasforoushan et al. 2019; McPherson et al. 2018b, 2018a; Owen et al. 2017).

## Data Availability

All data produced in the present study are available upon reasonable request to the authors.

## 6 Author Contributions

LMM and JPAD conceived and designed the research. LMM acquired and analyzed the data and prepared the initial manuscript. LMM and JPAD interpreted the data, edited and finalized the submitted manuscript, and agreed to be accountable for all aspects of the work.

## 8 Acknowledgments

The authors thank Carolina Carmona, PT, DPT for assistance with participant recruitment and screening and Becca Covode, B.S. for assistance with data collection.

